# Improved income status increased obesity and decreased anemia risk compared to high parity and low-income in pregnant south Ghanaian women: Analysis of hospital-acquired data

**DOI:** 10.1101/2022.12.19.22283683

**Authors:** Thomas Kwasi Awuni, Matsui Mitsuaki, Basma Ellahi, Francis Bruno Zotor

**Affiliations:** Ghana Health Service, Effutu Municipal Health Directorate, Ghana; School of Tropical Medicine and Global Health, Nagasaki University, Japan; Faculty of Health and Social Care, University of Chester, Chester, UK; Department of Family and Community Health, University of Health and Allied Science, Ghana

**Keywords:** Women, pregnant, overweight, obesity, income, anemia, hospital, Ghana

## Abstract

**Background:** Women of reproductive age (WIFA, 15-49 years) in Ghana are experiencing an upsurge in excess body mass index (BMI) and anemia (low hemoglobin concentration (Hb of ≤ 10.9 g/dl)). Regions of Southern Ghana (Central, Eastern, and Volta) are recognized to have high rates of adolescent pregnancies, and is known to be associated with high risk of anemia. However, little is known for pregnant South Ghanaian women. This study aimed to examine anthropometry – maternal stature and early pregnancy BMI – and socio-demographic measures as independent risk factors for anemia in antenatal women in Southern Ghana.

**Methodology:** Baseline data on anemia from healthy pregnant women (15-49 years; n = 1278) collected through antenatal attendance in an observational cohort study was included in the descriptive and logistic regression analysis using STATA (Stata Corp LLC [US]). Anemia (dependent) and the independent variables: high parity (≥5 children/woman); short stature (145≤155cm); underweight (BMI <18.5kg/m^2^), normal (BMI 18.5-24.9kg/m^2^), overweight (BMI 25-29.9kg/m^2^), and obesity (BMI >30 kg/m^2^) assessed with weight (kg) before the 20^th^ week of gestation divided by height (m^2^) were defined following standards. Wealth status was constructed using Principal Component Analysis of durable assets, housing characteristics, water/sanitation, and toilets facilities. All estimates were evaluated at the 5% significance level (p < 0.05).

**Results:** Of the sample, 45.85% had moderate/severe anemia with 9.67(±0.04) average hemoglobin (g/dl) concentration. Overweight/obesity was approximately (47%; overweight 29% vs obesity 18%), short stature (4.77%), and underweight (4.61%) in the 1278 women interviewed. Obesity was highest in the highest income women (26.24% (69/263)) in whom anemia decreased (34.78%, 80/230)) compared to highest prevalence of anemia in short stature (59.01%, 36/61), underweight (57.62%, 34/59), and overweight (44.68%, 164/367) women.

The risk of anemia increased with high parity (adjusted odds ratio; aOR 3.91; 95%CI: 1.79-8.52; p = 0.001); lowest income quintile (aOR:2.10; 95%CI: 1.35-3.26; p = 0.001); second income quintile (aOR:1.52; 95%CI: 1.01-2.30 p = 0.045); being Ewe ethnicity (aOR:2.09; 95%CI: 1.35-3.24; p = 0.001); and Akan (aOR:1.79; 95%CI: 1.16-2.74; p = 0.008); while obesity reduced the risk (aOR:0.70; 95%CI: 0.50-0.99; p = 0.045).

**Conclusion:** Anemia in pregnancy and overweight, and obesity as double burden of malnutrition remain significantly high in pregnant South Ghanaian women. The AIP exceeded the WHO defined threshold and was particularly highest in women having high parity, short stature, and underweight. While high parity and income status increased overweight/obesity, obese women were more likely to have a reduced risk of anemia. However, across the ethnicity, high parity and low-income status posed significant risk of anemia in women. Further research examining the nexus between underweight or short stature versus high rates of anemia or overweight/obesity and income and anemia to better explain, and prevent the increasing trend of anemia in pregnant Ghanaian women is recommended.

## Introduction

Malnutrition in all its forms is affecting humanity worldwide. Women in fertility age (WIFA, 15-49 years) are particularly vulnerable to overweight and obesity (excess BMI of ≤ 25kg/m^2^) and anemia (low hemoglobin concentration, Hb of ≤ 10.9/dl) in pregnancy (AIP). This is widely recognized as the overlooked paradox of triple burden of malnutrition [1–3]. Researchers posit that because societies have evolved embracing unhealthy lifestyles in addition to multiple childbearing practices or short inter-pregnancy intervals, nutritional stores of women often deplete alongside postpartum weight retention, thereby increasing the chances of overweight/obesity and AIP [1,4–6]. This often results in bleeding during delivery and other serious obstetric complications including low birth weight (less than 2.5kg) which in turn predicts many adult diseases [78].

Anemia in pregnancy and excess BMI varies by ethnicity and geographical locations and is endemic (more than 40%) in most Lower-Middle Income Countries (LMICs) [3,8–10]. Data from 46 LMICs indicates 40.4% of WIFA is anemic with pregnant women having 14% increased risk than non-pregnant women whilst those in the third trimester have 55% increased risk of anemia [3]. This is consistent with the World Health Organization (WHO) global statistics of anemia in WIFA (40%) and 42% in children less than 5 years [3]. In contrast, some LMICs such as Ethiopia and Tanzania reported a moderate level of anemia [11–13].

Anemia affects up to 42% of WIFA in Ghana more than the WHO defined 40% threshold at which mass iron supplementation is recommended in anemia endemic country. This record of anemia prevalence is also more than the global record and is been consistently high among WIFA across Northern, Middle and Southern zones of Ghana [3,14,15]. Also in Ghana, this prevalence is much more higher (48%) among adolescents, whose pregnancy represents 22.1%, 21.1%, and 17% in Southern Ghana (Volta, Central, and Eastern, the study regions respectively) compared to 14% known nationally, and pregnant women (45%) [14,15]. Overweight/obesity continues to soar in pregnant women (nearly 41 to 52% in 2008 and 2014 respectively) and among WIFA, 6% are estimated to be underweight, and 50% are overweight/obese [9,16,17]. Being overweight or obese potentially increases risk of iron deficiency (ID) due to low-grade inflammation which in turn elevates hepcidin production and subsequently, decreases iron efflux into circulation [2,18]. This reason was not linked to hemoglobin concentration and excess BMI. Likewise, underweight (BMI of < 18.5kg/m^2^) and short maternal stature have been shown to increase risk of anemia in pregnancy [2,19,20].

Anemia in pregnancy is associated with nutritional deficiencies, with ID accounting for 60% of severe and nearly 50% of moderate anemia cases worldwide.^2^ Similarly, in Ghana, ID underlies half of the anemia cases occurring in WIFA [14,16]. AIP is also common when women do not use birth control measures and have high parity, having life children of 5 or more alongside low-income and educational levels [5,6,11,15,21–24]. Women with high parity/fertility are most often poor due to the fact that they have more children to care for and cannot afford higher education with consequences for unemployment and a higher dependency ratio [11,15,21,23,25]. Thus, having more children increases household and family size with less opportunity for education, employment, income, and eventual poor maternal health [11,21,23,24].

Frequent childbearing or short interpregnancy intervals increase nutrient depletion, and this is particularly worse when nutrient inadequacy prolongs in the reproductive cycle, from conception through pregnancy and parturition [56]. Women have higher chances of experiencing hemorrhage during childbirth and depleting nutritional stores over time. Pregnancies result in an increased ID and anemia. Contrarily, others have retained postpartum weight [26,27].

Antenatal clinics (ANC) provide a system by which anemia and excess gestational weight gain as nutritional and birthing problems are addressed [22]. In Ghana, despite 87% of ANC attendees having attended at least four times during pregnancy, addressing anemia and preventing unhealthy weight gain during pregnancy lag behind ANC appointments [14–16]. Additionally, women empowerment programs and improvement in education have had little or no significant impact on childbearing age and parity or fertility rate. Total fertility in Ghana assessed as number of children *per* woman shows a medium-fertility level of 3.82 children *per* woman. This is a 1.72 increase above the required replacement-level fertility (2.1 children/woman) alongside a rising teen pregnancy rate [14,21,28]. Furthermore, a United Nations population statistics on birth control measures indicate Ghana is among African countries with low contraceptive use; median coverage of any method (22.2%), modern method (20.3%), and unmet need (34%) whilst only 36.2% had demand met [29]. These data are scary as it presupposed that a higher fertility/parity or a population increase, and associated risks of anemia in the WIFA population is eminent in Ghana.

A high parity to high BMI of ≥ 25kg/m^2^ association is common and further enhances risk of anemia in pregnant women. Such evidence is not well documented in Ghana where the double burden of malnutrition in pregnant women is reportedly high [9,14,17]. Further, it is unclear whether socioeconomic status, and anthropometric measures of above or below normal BMI and small maternal stature influences high levels of anemia in Ghanaian women. This study examined anthropometry – early pregnancy BMI and maternal stature – and socio-demographic measures as predisposition factors of AIP in women attending routine antenatal care in Southern Ghana (Central, Eastern, and Volta regions). This study is vital to know the current trend of AIP from these regions where the most available National representative data revealed a higher rates of adolescent pregnancy, a putative risk factor for anemia [14].

## Methodology

### Study design, population and sampling technique, sample size

Baseline data involving healthy pregnant women collected through antenatal attendance from an observational cohort study design conducted between October 2018 and May 2019 was included in this secondary data analysis. Women recruited were aged 15 to 49 years and were between 6 and 42 weeks of gestation. The gestational age was confirmed based on the reported last menstrual period cum ultrasound scan date [30].

Women with underlying conditions like Human Immune Deficiency Syndrome (HIV) or tuberculosis (TB) with potential to compromise nutritional status were excluded from the study. This was assessed through a review of their ANC booklet and through direct reported data from physician-diagnosed health conditions that could affect nutritional health. Participants who did not meet this defined criterion were excluded from the study.

A multi-stage sampling method was deployed to recruit the participants. The three regions as well as health facilities – a teaching hospital, three municipal hospitals, and two polyclinics – were purposely selected based on the documented evidence of adolescent pregnancy (Volta 22%, Central 21%, and Eastern 17%) [14,15]. This sampling method is vital to study a lager sample of women in LMICs for the explanation of the relationship between overweight/obesity and anemia [31]. The present study purposely included the health institutions for participants recruitment due to the fact that ANC attendance was high and the availability of medical doctor(s) or midwives with either basic essential obstetric or comprehensive care (BEmOC/EmOC), or both. This avoided loss to follow up (in the original study) as women were not referred elsewhere for delivery, as essential services that would be needed in the time of delivery were available.

The eligible participants were randomly recruited. Women were asked to pick a folded piece of paper upon entering the facility with ‘yes’ or ‘no’ written on. Those who picked ‘yes’ were interviewed after having understood the study procedures and willingly consented.

### Sample size

A sample size of 1278 was estimated using STATA (Stata Corp LLC [US]) power analysis command. The study was powered (80%) to detect 1.4 odds of association between maternal MUAC (of less than 23 cm) assuming prevalence (28%) of LBW infants [32]. There was a 10% adjustment to offset the effect of non-response rate, and as the minimum obstetric complication to be detected. The baseline data from the study was re-examined in this analysis to explain the influence of BMI, stature, and socio-demographic on AIP.

### Data collection

Trained nutrition officers and nurses employed face-to-face interviews and medical record review (ANC record booklet and attendance registers) to obtain data using a pre-tested semi-structured questionnaire. The questionnaire was adapted from a published document and piloted for necessary amendments before field implementation [14]. This ensured validity and reliability of the data analyzed. This process of data collection was performed whilst the participants were waiting to attend appointments with providers/midwives.

Primary data collected at baseline included economic status, and Hb (g/dl) concentration measured with a calibrated Hb meter (URIT Medical Electronic Co. Ltd, D-7, High-tech zone, China) through a standard procedure by laboratory technicians. The economic status assessment data was consistent with that used by national health surveys in Ghana and included a) *durable assets*: color television, radio set, refrigerator, mobile phone, car, or motorbike, gas, or electric stove, washing machine, electricity, computer, cabinet, internet access, and bed; b) *housing characteristics*: natural floor, cement floor, tiles, woolen carpet, rubber carpet, and roofing system; c) *water and sanitation*: pipe water, tube well, or borehole, surface water, sachet, or bottled water; and d) *toilet facility:* flush toilet, pit latrine, no toilet facility, and open defecation [14].

Retrospective data extracted from maternal and laboratory record books included maternal age, civil status, education, occupation, ethnicity, gravida, defined as the number times that a woman has been pregnant, parity, and sickling status. Anthropometric data of height and weight (kg) before the 20^th^ week of gestation was extracted from the records.

Training and monitoring of data collectors in addition to a field guide developed following standard protocol all of which allowed quality assurance of data collection and compliance to specified standards.

### Exposure and outcome variables

AIP was defined as low Hb (≤10.9g/dl) concentration including severe, moderate, and mild regardless of the gestation age. Those with adequate Hb concentrations of ≥11g/dl were classified as non-anemic [33].

Wealth index was estimated from asset data using Principal Components Analysis (PCA) method. This index measures inequalities in household characteristics as a proxy of women’s health services usage and eventual outcomes [14]. The binary (‘1’ as having the asset and ‘0’ not having the asset) data analyses gave a household wealth index score. Each household score was ranked into five equal groups, each reflecting 20% of the population. The resultant wealth quintiles were defined from the lowest to the highest quintile [14].

High parity/fertility level was defined as pregnant women having at least 5 life children at the time of the interview [23,28].

Anthropometry data included in this study were maternal stature and early pregnancy BMI. Maternal stature variable was grouped following standard as very short (<145 cm), short (145 to <150 cm), average (150 to <155cm), and standard (≥ 155cm) [19]. Early pregnancy BMI was computed using maternal weight (kg) before the 20^th^ week of gestation as the baseline weight divided by height in meters square (m^2^). The resultant BMI was further categorized into underweight (BMI of <18.5 kg/m^2^), normal (BMI of 18.5 to 24.9kg/m^2^), overweight (BMI of 25 to 29.9 kg/m^2^), and obese (BMI of > 30 kg/m^2^) [34].

### Data management, descriptive and outcome statistics

Data was collated in hard copy, cleaned, and double entered into Epidata software (version 4.6.0, http://www.epidata.dk/download.php, December 3, 2018). This was exported into STATA (Stata Corp LLC [US]) for analysis. Analyzed data was safely kept in locked facilities as hard copies and soft copies were password protected.

Descriptive analysis explored the prevalence of anemia in association with socioeconomic, maternal stature, and early pregnancy BMI variables as independent risk factors for AIP. The results were presented as percent distributions of Hb (g/dl) prevalence, socio-demographic factors, maternal stature, and BMI. The outcome analysis explored the relationship between the dependent and independent variables. Hb (g/dl) level was grouped into a binary variable, coded as ‘1’ if the respondent had anemia and ‘0’ if otherwise. Similarly, regarding wealth quintiles, the highest quintile was held as the reference group and primiparous as reference for the parity level.

Unadjusted univariable analysis between independent (socio-demographic and parity characteristics, maternal stature, and BMI) and the outcome/dependent (Hb status) variable were examined. Logistic regression with a backward selection procedure was implemented to develop the best fit multivariable model of associations between independent and the outcome variables. Variables were included in the multivariable model if loglikelihood ratio test (LRT) p-values were <0.05. Robust standard errors were calculated, using STATA ‘vce(robust)’ sub-command, to adjust for any correlation of the model errors due to the sampling procedure, assuming women who visited selected facilities shared similar covariates. Potential covariates retained in the final model included age, education, ethnicity, wealth status, gravid, parity, BMI, and Hb (g/dl).

### Ethics consideration

The study was approved by the Research Ethics Committee of the University of Health and Allied Science, Ghana, with the reference number; UHAS-REC A.2 [7], 18-19. Each selected health facility granted written permission for use of the study site. Each eligible woman who willingly volunteered to be involved provided written informed consent for enrollment after understanding the study details including potential benefits and harm, data collection procedures, data storage and protection, and anonymity as spelt out in the participants information sheet.

## Results

### Socioeconomic characteristics and anemia status

**Table 1** describes the background characteristics and anemia status of the respondents. Participants were 28.07 ±6.37 years, on average and teenage mothers (15-19 years) had the highest level of anemia (60.74% (82/135)). However, this consistently decreased for women aged to 41.82% (87/208) in the 35 and above age group with evidence of differences within group. Overall, the prevalence of AIP ((Hb ≤ 10.9 g/dl) was 45.85% (586/1278) with an average Hb (g/dl) level of 9.67 (±0.04) in anemic women, an indication of moderate anemia. Women in the 2^nd^ trimester constituted more than half (55.39% (708/1278)) of the study participants in whom 47.31% (335/708) were anemic.

**Table 1.** Sociodemographic characteristics and anemia status of antenatal mothers, southern Ghana.

| Variable | Anemia status |  |  | <sup>1</sup>p value |
| --- | --- | --- | --- | --- |
| | Not anaemic<br>(Hb ≥ 11 g/dl)<br>n = 692 (54.14) | Anaemic<br>(Hb ≤ 10.9 g/dl)<br>n = 586 (45.85) | Total<br>N = 1278 (100%) | |
| **Mean (SD) Hb level** | 12.20 (± 0.04) | 9.67 (± 0.04) | 11.04 (± 1.65) | |
| **Age (years, mean SD)** | 28.70 (± 0.22) | 27.33 (± 0.27) | 28.07 (± 6.37) | |
| 15-19 Adolescents | 53 (39.25) | 82 (60.74) | 135 (10.56) | 0.001 |
| 20-29 Young adult | 318 (53.08) | 281 (46.91) | 599 (46.87) |  |
| 30-34 Middle aged adults | 200 (59.52) | 136 (40.47) | 336 (26.29) |  |
| 35 and above Adults | 121 (58.17) | 87 (41.82) | 208 (16.27) |  |
| **Mothers' trimester** |  |  |  | 0.07 |
| 1<sup>st</sup> Trimester | 22 (73.33) | 8 (26.66) | 30 (2.34) |  |
| 2<sup>nd</sup> Trimester | 373 (52.68) | 335 (47.31) | 708 (55.39) |  |
| 3<sup>rd</sup> Trimester | 297 (55.00) | 243 (45.00) | 540 (42.25) |  |
| **Mothers' region of residency** |  |  |  | <0.001 |
| Central region | 251 (48.54) | 266 (51.45) | 517 (40.45) |  |
| Eastern region | 132 (70.58) | 55 (29.41) | 187 (14.63) |  |
| Volta region | 309 (53.83) | 265 (46.16) | 574 (44.91) |  |
| **Mother's ethnicity** |  |  |  | 0.08 |
| Akan | 315 (52.67) | 283 (47.32) | 598 (46.79) |  |
| Ewe | 296 (53.62) | 256 (46.37) | 552 (43.19) |  |
| Other (Ga/Northerners) | 81 (63.28) | 47 (36.71) | 128 (10.01) |  |
| **Mother's civil status** |  |  |  | 0.010 |
| Married | 544 (56.49) | 419 (43.50) | 963 (75.35) |  |
| In relationship | 105 (45.06) | 128 (54.93) | 233 (18.23) |  |
| Never married | 39 (52.00) | 36 (48.00) | 75 (5.86) |  |
| Divorced/widowed | 4 (57.14) | 3 (42.85) | 7 (0.54) |  |
| **Mother's educational status** |  |  |  | 0.001 |
| No formal/primary | 92 (46.46) | 106 (53.53) | 198 (15.49) |  |
| Junior High School | 355 (50.85) | 343 (49.14) | 698 (54.61) |  |
| Senior High School | 139 (61.23) | 88 (38.76) | 227 (17.76) |  |
| Tertiary | 106 (68.38) | 49 (31.61) | 155 (12.12) |  |
| **Mother's occupation** |  |  |  | <0.001 |
| Artisan | 119 (50.63) | 116 (49.36) | 235 (18.38) |  |
| Farmer | 33 (47.82) | 36 (52.17) | 69 (5.39) |  |
| Trading | 283 (56.26) | 220 (43.73) | 503 (39.35) |  |
| Employed (public/private) | 142 (65.74) | 74 (34.25) | 216 (16.90) |  |
| No defined job | 115 (45.09) | 140 (54.90) | 255 (19.95) |  |
| **Gravida of the mother** |  |  |  | 0.002 |
| 1<sup>st</sup> Pregnancy | 148 (46.39) | 171 (53.60) | 319 (24.96) |  |
| 2-4<sup>th</sup> Pregnancy | 442 (57.92) | 321 (42.07) | 763 (59.70) |  |
| ≥ 5<sup>th</sup> Pregnancies | 102 (52.04) | 94 (47.95) | 196 (15.33) |  |
| **Parity of the mother** |  |  |  | <0.001 |
| Primiparous (1<sup>st</sup> child) | 366 (52.66) | 329 (47.33) | 695 (54.38) |  |
| Multiparous (2-4<sup>th</sup> child) | 311 (58.45) | 221 (41.54) | 532 (41.62) |  |
| Grand multiparous (≥ 5 children) | 15 (29.41) | 36 (70.58) | 51 (3.99) |  |
| **Wealth quintiles** |  |  |  | <0.001 |
| Lowest | 99 (39.60) | 151 (60.40) | 250 (19.56) |  |
| Second | 125 (50.40) | 123 (49.59) | 248 (19.40) |  |
| Middle | 140 (54.68) | 116 (45.31) | 256 (20.03) |  |
| Fourth | 155 (59.38) | 106 (40.61) | 261 (20.42) |  |
| Highest | 173 (65.77) | 90 (34.22) | 263 (20.57) |  |
| **Sickle cell anemia** |  |  |  |  |
| Positive | 15 (50.00) | 15 (50.00) | 30 (2.34) |  |
| Negative | 677 (54.24) | 571 (45.75) | 1248 (97.65) |  |
| **Maternal stature (cm)** |  |  |  | 0.05 |
| Very short (< 145 cm) | 6 (50.00) | 6 (50.00) | 12 (0.93) |  |
| Short (145 to < 150 cm) | 25 (40.98) | 36 (59.01) | 61 (4.77) |  |
| Average (150 to < 155 cm) | 95 (48.96) | 99 (51.03) | 194 (15.17) |  |
| Normal ($\geq 155$ cm) | 566 (55.98) | 445 (44.01) | 1011 (79.10) | |
| **Mothers' pre-pregnancy BMI** |  |  |  | <0.001 |
| Undernutrition (< 18.5 kg/m<sup>2</sup>) | 25 (42.37) | 34 (57.62) | 59 (4.61) |  |
| Normal ($\geq 18.5$ to 24.9 kg/m<sup>2</sup>) | 314 (50.48) | 308 (49.51) | 622 (48.66) | |
| Overweight ($\geq 25$ to 29.9 kg/m<sup>2</sup>) | 203 (55.31) | 164 (44.68) | 367 (28.71) | |
| Obese ($\geq 30$ kg/m<sup>2</sup>) | 150 (65.21) | 80 (34.78) | 230 (17.99) | |

N: total sample; n: sub-sample size; Hb: haemoglobin; g/dl: grams *per* deciliter; SD: standard deviation; ≥ : greater than or equal to; %: per cent; BMI: body mass index; kg/m<sup>2</sup>: kilogram per meter square; cm: centimetre; <sup>1</sup>Pearson's chi square test.

The greatest proportions of the women were recruited from the municipal and teaching hospitals in Volta region (44.91% (574/1278)) with the least being from the municipal hospital in the Eastern region (14.63% (187/1278)). Anemia was disproportionally higher in Central (51.45% (266/517)) followed by Volta (46.16% (265/574)), and Eastern (29.41% (55/187)). Anemia was highest in Akan ethnicity (47.32% (283/598)), followed by Ewes (46.37% (256/522)) while Ga-Dangme and Northerners together had the least (36.71 % (47/128)). This was consistent with the study population pattern; Akan (46.79% (598/1278%), Ewes (43.19% (552/1278)), and the minority were Ga-Dangme and Northerners (10.01% (128/1278)).

Low income and primary/no formal educational levels were associated with increased prevalence of AIP. Of the 1278 women Hb(g/dl) data included in the analyses, 60.40% (151/250) and 49.59% (123/248) of those in the lowest and second income quintile respectively were anemic compared to those in the highest income quintile (34.22% (90/263)). More than half (54.61% (698/1278)) of the total respondents had completed junior high school compared to those who had attained a tertiary degree (12.12% (155/1278)). Those with no formal/primary education had highest level of AIP (53.33% (106/198)), but the number decreased among those who attained tertiary grade (31.61% (49/155)). Anemia was highest in women who had no formal jobs (54.90% (140/255)), followed by farmers (52.17% (36/69)), but this decreased to 34.25% (74/216) in private/public sector employees.

More than half (54.38% (695/1278)) of the respondents were primiparous (having a child for the first time) with around 4% (51/1278) of them in the high parity (having at least five children) among whom a highest proportion (70.58% (36/51)) were anemic.

Data on maternal stature shows that those with normal height and anemic were fewer, meanwhile, the number increased in short stature women (59.01% (36/61)), though this cohort forms just about 5% (61/1278) of the study population. This dropped by nearly 8% in those within the average height (51.03% (99/194) while declining further by 15% in those attaining a normal height (44.01% (445/1011)).

Early pregnancy BMI assessment indicate high levels of overweight/obesity ((46.7%; 597/1278; overweight (28.71% (367/1278)) *vs* obesity (17.99% (230/1278)). Anemia was highest in underweight (57.62% (34/59)) though this cohort represented only about 5% (59/1278) of the study population. This consistently decreased by 8% in the normal BMI category (49.51% (308/622) even though this group forms the highest proportion of the interviewees (48.66% (622/1278)). Nearly 13% reduction was evident in overweight (44.68% (164/367)) compared to almost 23% decline in the obese women (34.78% (80/230) respectively from the underweight category (**Table 1**). In **Figure 1** early pregnancy BMI category varies by women’s parity level. Obesity increased by 8.78% from 13.95% (97/695) in primiparous women to 22.74% (121/532) in multiparous (2-3 children/woman) and increased further by 9.58% to 23.53% (12/51) in high parity women.

**Fig 1:**
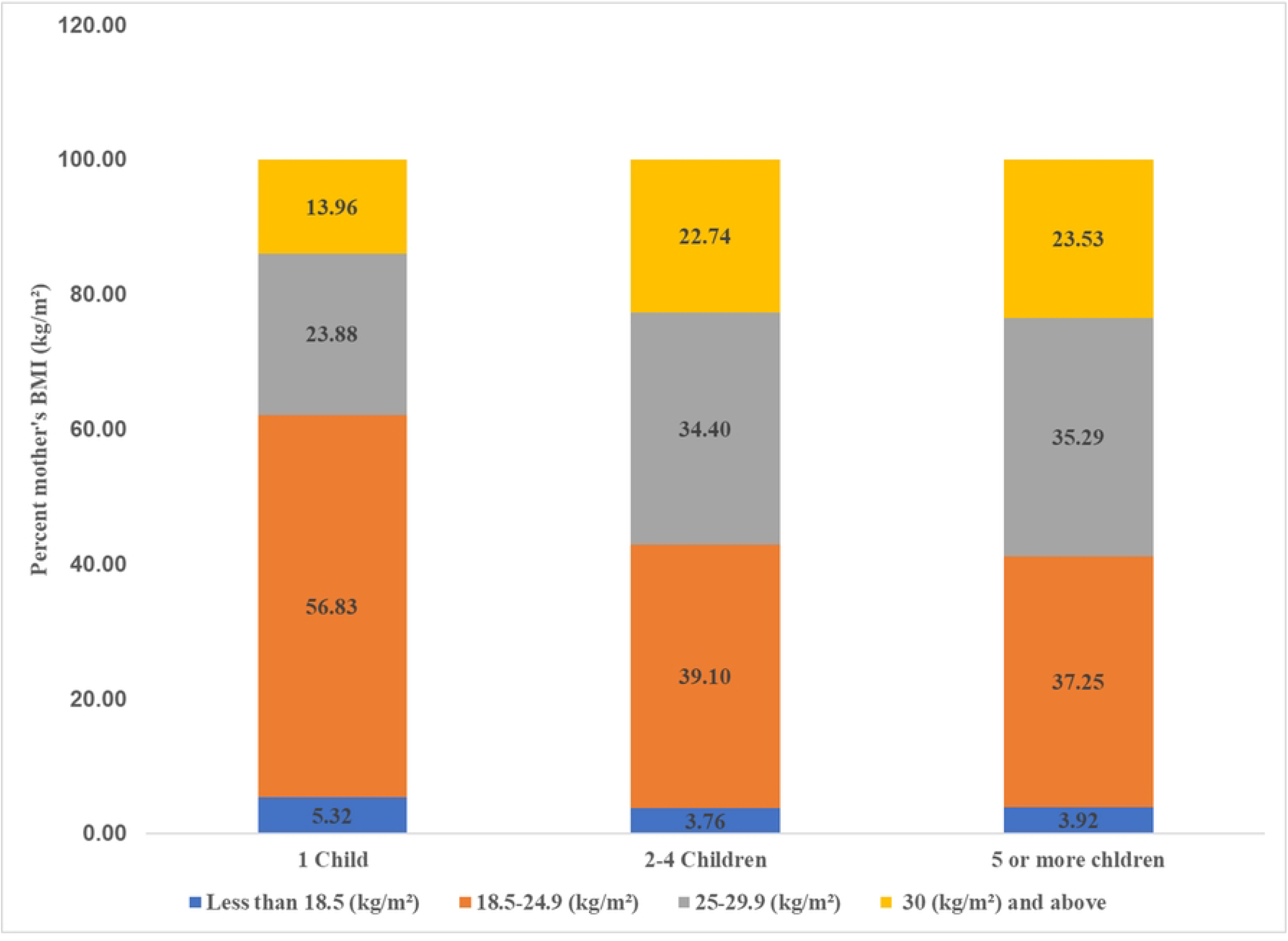
Percent distribution of early pregnancy BMI by mother’s parity*. (*Primiparous, n = 695; Multiparous, n = 532; Grand multiparous, n = 51)

Figure 2. reveals that early pregnancy BMI category varied by women’s income level. Healthy weight gain was highest (63.6% (159/250)) in the lowest income women, but this decreased by nearly 24% to 39.92% (105/263) in the highest income women in whom obesity was highest (26.24% (69/263)). Obesity was lowest (9.20%, 23/250)) in the lowest income women. So, the wealthier the women were the higher the rates of excess BMI and vice-versa.

**Fig 2:**
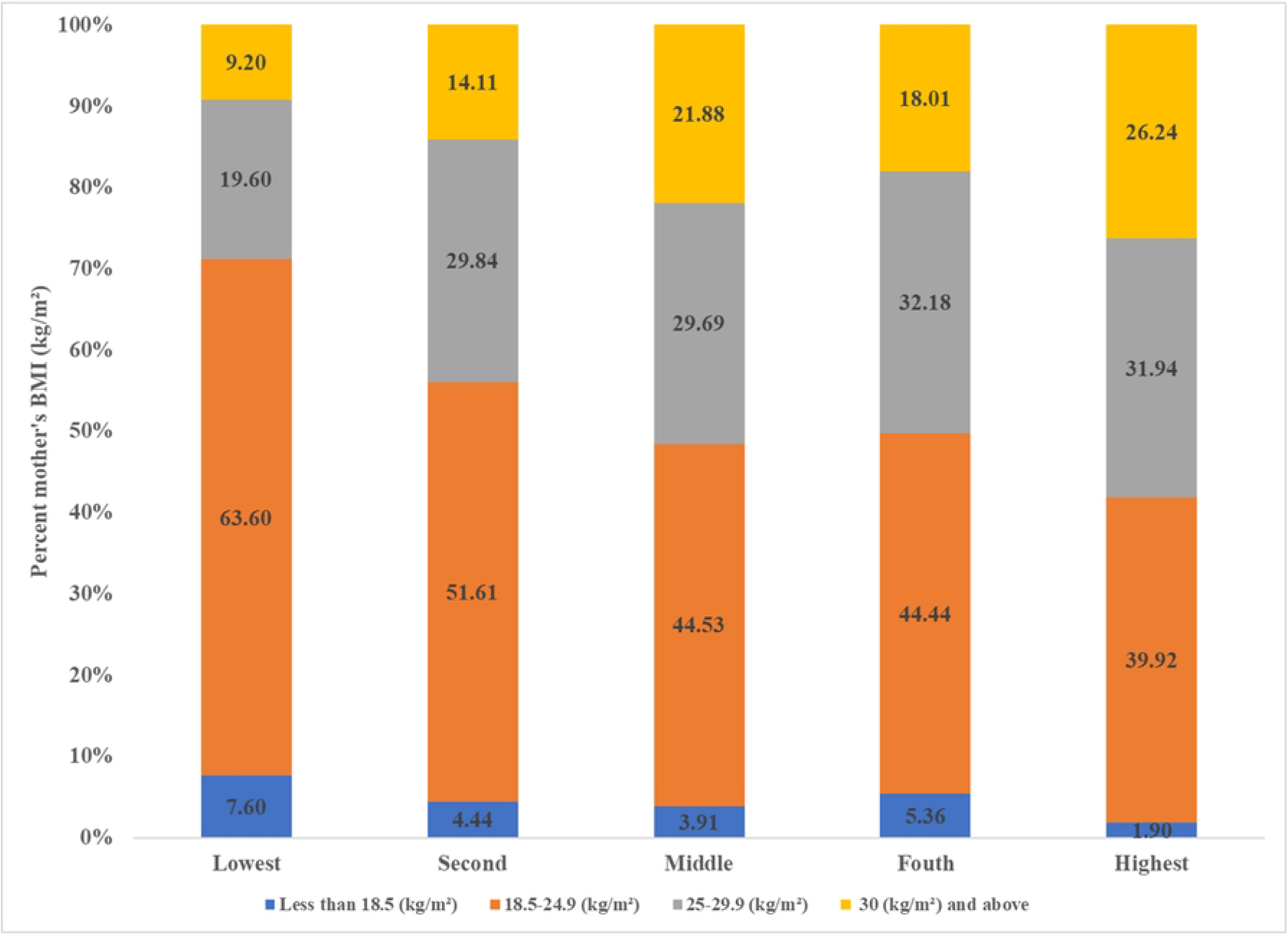
Percent distribution of early pregnancy BMI by mother’s income level*. (*Lowest, n = 250; Second, n = 248; Middle, n = 256; Fourth, n = 261; Highest, n = 263)

### Results of anemia in pregnancy and associated risk factors

**Table 2** outlines the odds ratio (OR) and adjusted odds ratio (aOR) of factors associated with AIP. Being from an Ewe ethnicity was associated with high risk of anemia (OR = 1.49; 95% CI: 1.00 to 2.21; p = 0.048) and (aOR = 2.09; 95% CI: 1.35 to 3.24; p 356 = 0.001), and (OR = 1.54, 95% CI: 1.04 to 2.29; p = 0.029), and (aOR = 1.79; 95% CI: 1.16 to 2.74; p = 0.008) if an Akan ethnicity.

**Table 2.** Logistic regression of socio-demographic characteristics and anemia status of antenatal mothers, southern Ghana.

| Variable | Anemia status |  | Unadjusted and Adjusted odds ratio |  |  |  |
| --- | --- | --- | --- | --- | --- | --- |
| | Not anaemic<br>(Hb ≥ 11 g/dl)<br>n = 692<br>(54.14%) | Anaemic<br>(Hb < 11.0 g/dl)<br>n = 586 (45. 8%) | Unadjusted<br>OR (95% CI) | P-value | Adjusted<br>aOR (95% CI) | P-value |
| **Age (years, mean SD; 28.04 (± 6.38))** |  |  |  | **<0.001** |  |  |
| 15-19 Adolescents | 53 (39.25) | 82 (60.74) | 1.75 (1.19, 2.56) | 0.004 | 1.16 (0.71, 1.90) | 0.547 |
| 20-29 Young adult | 318 (53.08) | 281 (46.91) | Reference |  | Reference |  |
| 30-34 Middle aged adults | 200 (59.52) | 136 (40.47) | 0.76 (0.58, 1.00) | 0.058 | 0.88 (0.64, 1.20) | 0.417 |
| 35 and above Adults | 121 (58.17) | 87 (41.82) | 0.81 (0.58, 1.11) | 0.205 | 0.76 (0.51, 1.13) | 0.185 |
| **Mother's ethnicity** |  |  |  | **0.08** |  |  |
| Akan | 315 (52.67) | 283 (47.32) | 1.54 (1.04, 2.29) | 0.029 | 1.79 (1.16, 2.74) | 0.008 |
| Ewe | 296 (53.62) | 256 (46.37) | 1.49 (1.00, 2.21) | 0.048 | 2.09 (1.35, 3.24) | 0.001 |
| Other (Ga/Northerners) | 81 (63.28) | 47 (36.71) | Reference |  | Reference |  |
| **Mother's educational status** |  |  |  | **<0.001** |  |  |
| No formal/primary | 92 (46.46) | 106 (53.53) | 2.49 (1.60, 3.86) | <0.001 | 1.66 (0.92, 3.02) | 0.091 |
| Junior High School | 355 (50.85) | 343 (49.14) | 2.09 (1.44, 3.02) | <0.001 | 1.40 (0.83, 2.37) | 0.197 |
| Senior High School | 139 (61.23) | 88 (38.76) | 1.36 (0.88, 2.10) | 0.153 | 1.06 (0.63, 1.78) | 0.819 |
| Tertiary | 106 (68.38) | 49 (31.61) | Reference |  | Reference |  |
| **Mother's civil status** |  |  |  | **0.01** |  |  |
| Married | 544 (56.49) | 419 (43.50) | Reference |  | Reference |  |
| In relationship | 105 (45.06) | 128 (54.93) | 1.58 (1.18, 2.11) | 0.002 | 1.01 (0.71, 1.42) | 0.955 |
| Never married | 39 (52.00) | 36 (48.00) | 1.19 (0.74, 1.37) | 0.451 | 0.79 (0.45, 1.37) | 0.406 |
| Divorced/widowed | 4 (57.14) | 3 (42.85) | 0.97 (0.21, 4.37) | 0.972 | 0.96 (0.20, 4.63) | 0.969 |
| **Mother's occupation** |  |  |  | **<0.001** |  |  |
| Artisan | 119 (50.63) | 116 (49.36) | Reference |  |  |  |
| Farmer | 33 (47.82) | 36 (52.17) | 1.11(0.65, 1.91) | 0.68 | 0.74 (0.41, 1.35) | 0.333 |
| Trading | 283 (56.26) | 220 (43.73) | 0.79 (0.58, 1.08) | 0.153 | 0.83 (0.60, 1.16) | 0.288 |
| Employed (public/private) | 142 (65.74) | 74 (34.25) | 0.53 (0.36, 0.78) | 0.001 | 0.83 (0.51, 1.34) | 0.458 |
| No defined job | 115 (45.09) | 140 (54.90) | 1.24 (0.87, 1.78) | 0.22 | 1.02 (0.66, 1.56) | 0.922 |
| **Gravida of the mother** |  |  |  | 0.002 |  |  |
| 1<sup>st</sup> Pregnancy | 148 (46.39) | 171 (53.60) | Reference |  | Reference |  |
| 2-4<sup>th</sup> Pregnancy | 442 (57.92) | 321 (42.07) | 0.62 (0.48, 0.81) | 0.001 | 0.79 (0.56, 1.12) | 0.199 |
| ≥5 Pregnancy | 102 (52.04) | 94 (47.95) | 0.79 (0.55, 1.13) | 0.214 | 0.76 (0.44, 1.32) | 0.345 |
| **Parity of the mother** |  |  |  | <0.001 |  |  |
| Primiparous (1<sup>st</sup> child) | 366 (52.66) | 329 (47.33) | Reference |  | Reference |  |
| Multiparous (2-4<sup>th</sup> child) | 311 (58.45) | 221 (41.54) | 0.79 (0.62, 0.99) | 0.043 | 0.97 (0.71, 1.33) | 0.897 |
| Grand multiparous (≥ 5 children) | 15 (29.41) | 36 (70.58) | 2.66 (1.43, 4.96) | 0.002 | 3.91 (1.79, 8.52) | 0.001 |
| **Wealth quintiles** |  |  |  | <0.001 |  |  |
| Lowest | 99 (39.60) | 151 (60.40) | 2.93 (2.04, 4.19) | <0.001 | 2.10 (1.35, 3.26) | 0.001 |
| Second | 125 (84.45) | 123 (83.10) | 1.89 (1.32, 2.70) | <0.001 | 1.52 (1.01, 2.30) | 0.045 |
| Middle | 140 (54.68) | 116 (45.31) | 1.59 (1.11, 2.26) | 0.010 | 1.27 (0.85, 1.89) | 0.225 |
| Fourth | 155 (59.38) | 106 (40.61) | 1.31 (0.92, 1.87) | 0.131 | 1.09 (0.74, 1.61) | 0.632 |
| Highest | 173 (65.77) | 90 (34.22) | Reference |  | Reference |  |
| **Mothers' BMI (kg/m<sup>2</sup>)** |  |  |  | 0.004 |  |  |
| Undernutrition (< 18.5 kg/m<sup>2</sup>) | 25 (42.37) | 34 (57.62) | 1.38 (0.80, 2.37) | 0.235 | 1.31 (0.74, 2.32) | 0.343 |
| Normal (≥18.5.0 to 24.9 kg/m<sup>2</sup>) | 314 (50.48) | 308 (49.51) | Reference |  | Reference |  |
| Overweight (≥ 25 to 29.9 kg/m<sup>2</sup>) | 203 (55.31) | 164 (44.68) | 0.82 (0.63, 1.06) | 0.142 | 0.97 (0.73, 1.28) | 0.779 |
| Obese (≥ 30 kg/m<sup>2</sup>) | 150 (65.21) | 80 (34.78) | 0.54 (0.39, 0.74) | <0.001 | 0.70 (0.50, 0.99) | 0.045 |

N: total sample; n: sub-sample size; Hb: haemoglobin; g/dl: grams per deciliter; SD: standard deviation; ≥: greater than, or equal to; ≤: less than, or equal to; %: per cent; CI: confidence interval; BMI: body mass index; kg/m<sup>2</sup>: kilogram *per* meter square

The risk of anemia was highest in the high parity (5 children/women) (OR = 2.66; 95% CI: 1.43 to 4.96; p = 0.002) and (aOR = 3.91; 95% CI: 1.79 to 8.52; p = 0.001). Surprisingly, being obese was associated with 46% reduced risk of anemia (OR = 0.54; 95% CI: 0.39 to 0.74; p < 0.001) and 30% reduced risk (aOR = 0.70; 95% CI: 0.50 to 0.99; p = 0.045).

Whereas, neither gravida, education, or occupation showed no impact on AIP, the risk of AIP was highest, nearly 3 times (OR = 2.93; 95% CI: 2.04 to 4.19, p < 0.001) and aOR of 2.1 times (aOR = 2.10; 95% CI: 1.35 to 3.26; p = 0.001) among women in the lowest income quintile. Similarly, the second quintile group had (OR = 1.89, 95% CI: 1.32 to 2.70; p < 0.001) and (aOR = 1.52; 95% CI: 1.01 to 2.30; p = 0.045) risk of becoming anemic. The risk significantly disappeared as the women wealth status improved in the middle, fourth and the highest quintile.

## Discussion

High proportion of pregnant women interviewed at routine antenatal care in the selected regions of Southern Ghana were anemic and overweight, and obese reflecting the growing double burden of malnutrition in this population group. Statistics across the world indicate pregnant women or WIFA (15-49 years) are either anemic or overweight and obese, or both [1]. Most significantly, the anemia recorded was higher, than the WHO defined 40% threshold [8,14,15]. The present findings uniquely identified anemia as a micronutrient deficiency increasingly trending on a par with macronutrient malnutrition [4,15]. This is consistent with National surveys. In particular, the Ghana Demographic Health Survey data revealed a higher burden of maternal malnutrition, varying substantially by individual nutritional status, economic status, parity, and ethnicity [14,35,36]. These are paradoxically linked predisposition factors of malnutrition common to LMICs [1,4,37].

Anemia was significantly more common in high parity and low (lowest and second quintile) income women groups in whom close to two-third and more than half respectively had healthy weight (BMI of 18.5 to 24.9kg/m^2^) than highest income women among whom obesity increased. While overweight and obesity increased positively with increasing parity and high-income level, there was an inverse relationship between obesity and the rate of anemia. The high parity being proportionate to high level of anemia and overweight/obesity enhances the fact that child bearing practices increase nutrient depletion as well as promote blood loses and postpartum weight retention [5,6,11,23,26,27,38]. Though statistically insignificant, AIP was highest in short stature (145≤155cm) and underweight (BMI <18.5kg/m^2^) women than their counterparts of healthy weight (BMI of 18.5 to 24.9kg/m^2^) and compares with evidence, especially from rural India and the Maldives [2,20,39].

Despite the fact that the present analysis not including dietary habits as a determinant of malnutrition, it is possible the poor and undernourished (short and underweight) pregnant women were more anemic because of nutritional inadequacies in the diet whilst the high economic classified women indulged in high caloric diets with little or no physical activity resulting in higher levels of overweight/obesity [4]. This lifestyle lays the foundation for malnutrition to occur, a phenomenon commonly associated with nutrition transition where urbanization and modernization accompanies income and economic growth [4]. This further suggests perchance, women in poor income quintile consumed less caloric dense diets (which may be of a poorer nutritional quality but may not be nutrition secured and adequately utilize nutrients) and or undertook hard physical work compared to the higher income ladies thus are less likely to be overweight or obese, but were anemic.

Studies expound the excess BMI and iron deficiency (ID) and iron deficiency anemia (IDA) relationships [2,18,37]. Overweight/obesity prior to and during pregnancy imposed a higher ID and IDA burden on infant-mother-pair, and nearly doubles the risk of postpartum anemia in women [2,18,37,40]. A woman who is overweight or obese has increased risks of ID due to natural process avoiding human and pathogen competition for iron discussed earlier [2]. In contrast, there are inconclusive evidence about overweight/obesity and Hb (g/dl) concentration in women. The obese women in this study had 30% reduced risk of AIP consistent with other studies including one study involving obese Chinese women who had improved Hb(g/dl) concentration level [2,37,40–42]. The present study findings reasonably suggest that obesity protects against AIP in contrast to high parity which significantly underpin increased obesity level as well as increased anemia risk.

Evidently, having at least 5 children/women increase overweight/obesity risk and independently, predisposed women to nearly four-fold risk of anemia in this study. Though the number of women in this cohort was small (about 4%), the finding is similar to that of a study in Oman where approximately half of the women studied had high parity with a significantly higher incidence of AIP [23]. The same risk was not evident when analyzed according to the number of pregnancies, not live births in this study. Short interpregnancy interval, or high parity/fertility depletes nutrition reserves and thus, predisposes women to a higher risk of anemia [5,6,11]. Of note, while age at pregnancy had no significant relation with AIP in this study, the findings support evidence that increase childbearing influences nutrient depletions, a putative risk factor for anemia in the reproductive cycle [5,6,23].

The result also exemplified anemia as a condition of the poor and overweight/obesity as conditions of the affluents. Whereas, poverty was inversely associated with overweight/obesity in women, poverty and high parity were positively associated with increased risk of anemia. Evidently, when women’s economic status improved, the risk of anemia being more likely decreases while overweight and obesity increased. In other words, women in the lowest and second quintiles were more likely to have anemia than their counterparts in the highest income quintiles, but not overweight/obesity. It could be speculated that poorer women are more likely to have many children, which may push them into poverty and thereby they may be unable to afford healthy diets to prevent AIP. Indeed, having 5 or more children predisposed women to low income (poverty) and anemia, but not overweight and obesity in this study. Studies in part of Asia and Eastern Africa demonstrated that a lower average monthly income and high parity were vital modifiable risk factors for rising levels of AIP [12,43,44]. Conversely, birth control strategies and education reduced the risk of AIP [44,45]. These results underscore the significance of improving individuals or household income status whilst maintaining a replacement-fertility level, particularly among the poor, as recommended in other studies [8,10,11,21,44,45].

The direct association between high parity, poverty, and increased likelihood of AIP found in the present study was associated with frequent birthing experiences in other literatures [45]. For instance, whereas Oman reported a higher incidence of AIP in high parity women, analysis of Myanmar’s first Demographic and Health Survey data revealed elevated risks of AIP prevalence [46]. Understandably, high parity women have higher chances of experiencing hemorrhage during childbirth due to malnourishment associated with frequent pregnancies or short interpregnancy intervals [5,6,23]. Another plausible reason could be that having more children increases household/family size, which may result in women being unable to get adequate nutrition, thus explaining the apparent different risk for anemia in relation to the number of pregnancies, compared to live births and also the low income suggested in the present study.

A larger proportion of pregnant adolescents were anemic compared to the adults however the results were not statistically significant. This finding seems to contrast the findings that high parity, a factor associated with age, increases the risk of AIP [5,6,23,46]. Indeed, the result demonstrated that high parity resulted in an increased odd of AIP, but not in association with adulthood. The higher proportion of AIP in the adolescents could be attributed to their inability to meet daily nutrient requirements prior to pregnancy and thereby unable to meet the increased nutritional demand in pregnancy [15]. This may be due to the inability to sustain iron and hemoglobin levels during menstruation in adolescence.

Ethnicity was an unmodifiable risk factor for overweight/obesity and anemia in pregnant women across the study regions. The residents of the Central region, most of whom were of the Akan ethnic origin, had the highest proportion of AIP consistent with a recent study in the region [47]. The Ewes who are predominantly domiciled in the Volta region came second following Central regional record of anemia. However, women from Volta had the highest odds – twice the risk – of AIP among the ethnics compared to Akans who had about 1.8 times risk. However, obesity was higher (nearly one-fifth) in Central and Eastern respectively compared with about one-sixth in Volta (figure not shown). These levels of obesity and anemia in Volta region cast doubt that obesity reduces anemia risk. Nonetheless, elsewhere, in Saudi Arabia obese pregnant women, about three-fifths were anemic [48], whereas analysis of a National data from Ghana indicated below 10% co-occurrence of overweight/obesity and anemia in WIFA, [36] but more than half were shown to have co-occurrence of overweight and anemia [35].

The regional and ethnic variations in anemia and overweight/obesity found in Southern Ghana corroborate previous studies in the Middle and Northern parts of the country [8–10]. Elsewhere, secondary analysis of the Colombian National Nutrition Survey (ENSIN) 2010 data elucidated similar associations of high levels of AIP across geographic groups/ethnicities [49]. However, developing country like Ethiopia and Tanzania reported a moderate level of anemia in women whilst some high-income countries such as New Zealand, Australia, England, and Ireland, reported below three percent prevalence [11–13].

The established burden of anemia and overweight, and obesity in pregnant South Ghanaian women enhances the WHO recommendation for increase ANC attendance where women anthropometry, dietary counselling, and socioeconomic measurements are assessed before delivery [22]. And whereas obesity seems protective against AIP in this study in support of other studies, but contrarily to many other findings, it is important to deploy alternative diagnostic methods to further explain the rising trend of overweight/obesity and anemia and the causes in Ghanaian women [2,18,50]. For instance, measuring ID including detecting inflammation is supported in a WHO guideline where ferritin concentration is recommended for iron status assessment [50]. A review of overweight/obesity and risk of ID/anemia by Wawer et al. corroborates the WHO’s guideline [2]. Mwangi and colleagues have, however, cited uncertainties in the use of zinc protoporphyrin (ZPP) in addition to Hb concentration to define AIP [51]. Regardless of the findings, a further review has advocated for its use [51].

### Limitations and strengths

Cause and effect relationships are hard to determine since the data were drawn from an observational cohort study baseline data. Dietary habits and preventive services like iron supplementation, helminths, and infection prevention during pregnancy which are important risk factors were not studied, hence misclassification bias is likely. Further, the analysis also included baseline data of Hb(g/dl) concentration measurement that has several limitations in anemia estimation than iron status (for example, serum ferritin) assessment [50]. Additionally, the regions and the health facilities were conveniently selected and no urban-rural segregation could potentially bias the results. However, the method was recommended as necessary for LMICs [31].

Consequently, the researcher has made cautious interpretations of the results and deployed anthropometry data to determine nutritional effect on maternal anemia. The study also enhanced the reliability of data through the use of a validated tool by skilled professionals who had training on the study method and were provided with standard references for data collection. In addition, field investigators had assigned supervisors knowledgeable in nutritional and health surveys.

## Conclusion

Anemia in pregnancy and overweight, and obesity as a double burden of malnutrition remains significantly high in pregnant South Ghanaian women. The AIP exceeded the WHO defined threshold as a significant public health threat and was particularly highest in women having high parity, short stature, and underweight. While high parity and income status increased overweight/obesity, obese women were more likely to have a reduced risk of anemia. However, across the ethnicity, high parity and low-income status posed significant risk of anemia in women. As women with an improved income status were more likely at risk of overweight/obesity but less anemic, public health nutrition recommendations should focus on targeting these higher income women to ensure they focus on more healthy diets and physical activity to reduce excess BMI.

## Data Availability

All relevant data are within the manuscript and its Supporting Information files.

## Acknowledgments

The authors would like to acknowledge the following individuals and organizations for their role(s): Japanese Government, staff of Japanese Grant Aid for Human Resource Development, and Japan International Cooperation Agency (JICA) for funding my studies in Japan. Nagasaki University School of Tropical Medicine and Global Health (TMGH), Japan, for Student Research Support provided. Management and staff of various health facilities where the study was conducted. All field supervisors and research assistants whose contributions ensured collection of data.

Shafiq Siita, National Health Insurance, Upper East Regional Office, Ghana, for guidance on data management.

## Author contributions

Study conception TKA, MM, BE, FBZ, preliminary analysis and graphics TKA, MM, drafting the manuscript TKA, preparing results tables MM, manuscript critical review and finalization of manuscript for submission BE, FBZ.

## Supporting information

**S1 Dataset. This is the S1 File Title**.

## References

1. Launbo N, Davidsen E, Granich-Armenta A, et al. The overlooked paradox of the coexistence of overweight/obesity and anemia during pregnancy. Nutrition. 2022;99–100. doi:10.1016/j.nut.2022.111650

2. Wawer AA, Hodyl NA, Fairweather-tait S, Froessler B. Are pregnant women who are living with overweight or obesity at greater risk of developing iron deficiency/anaemia? Nutrients. 2021;13,(1572). doi:https://doi.org/10.3390/nu13051572

3. Sun J, Wu H, Zhao M, Magnussen CG, Xi B. Prevalence and changes of anemia among young children and women in 47 low-and middle-income countries, 2000-2018. eClinicalMedicine. 2021;41. doi:10.1016/j.eclinm.2021.101136

4. WHO. The Double Burden of Malnutrition. Policy Brief. Vol 21.; 2017. doi:10.1111/j.1745-6584.1983.tb00740.x

5. Winkvist A, Rasmussen KM, Jean-Pierre Habicht. A New definition of maternal depletion syndrome. Am J Public Heal. Published online 1992:691–694.

6. King JC. The risk of maternal nutritional depletion and poor outcomes increases in early or closely spaced pregnancies. J Nutr. 2003;133(5 SUPPL. 1):1732–1736. doi:10.1093/jn/133.5.1732s

7. Figueiredo ACMG, Gomes-Filho IS, Batista JET, et al. Maternal anemia and birth weight: A prospective cohort study. PLoS One. 2019;14(3):e0212817. doi:10.1371/journal.pone.0212817

8. Ayensu J, Annan R, Lutterodt H, Edusei A, Peng LS. Prevalence of anaemia and low intake of dietary nutrients in pregnant women living in rural and urban areas in the Ashanti region of Ghana. PLoS One. 2020;15(1):1–15. doi:10.1371/journal.pone.0226026

9. Ofori-asenso R, Agyeman AA, Laar A, Boateng D. Overweight and obesity epidemic in Ghana — a systematic review and meta-analysis. BMC Public Health. Published online 2016. doi:10.1186/s12889-016-3901-4

10. Ahenkorah B, Nsiah K, Baffoe P. Sociodemographic and Obstetric Characteristics of Anaemic Pregnant Women Attending Antenatal Clinic in Bolgatanga Regional Hospital. Scientifica (Cairo). 2016;2016:13–16. doi:10.1155/2016/4687342

11. Bekele A, Tilahun M, Mekuria A. Prevalence of Anemia and Its Associated Factors among Pregnant Women Attending Antenatal Care in Health Institutions of Arba Minch Town, Gamo Gofa Zone, Ethiopia: A Cross-Sectional Study. Anemia. 2016;2016:1–9. doi:10.1155/2016/1073192

12. Stephen G, Mgongo M, Hussein Hashim T, Katanga J, Stray-Pedersen B, Msuya SE. Anaemia in Pregnancy: Prevalence, Risk Factors, and Adverse Perinatal Outcomes in Northern Tanzania. Anemia. 2018;2018. doi:10.1155/2018/1846280

13. Masukume G, Khashan AS, Kenny LC, Baker PN, Nelson G. Risk factors and birth outcomes of anaemia in early pregnancy in a nulliparous cohort. PLoS One. 2015;10(4):1–15. doi:10.1371/journal.pone.0122729

14. GSS GHS and ICF International. Ghana Demographic and Health Survey 2014.; 2015.

15. WHO. Guideline: Daily Iron Supplementation in Adult Women and Adolescent Girls.; 2016.

16. University of Ghana, GroundWork, University of Wisconsin-Madison, KEMRI-Wellcome Trust U. Ghana Micronutrient Survey 2017.; 2017.

17. Ghana Statistical Service Ghana Health Service (GSS) ICF and International. Ghana Demographic and Health Survey 2009. Accra, Ghana: GSS, GHS, and ICF Macro.; 2009.

18. Bodnar LM, Siega-Riz AM, Cogswell ME. High prepregnancy BMI increases the risk of postpartum anemia. Obes Res. 2004;12(6):941–948. doi:10.1038/oby.2004.115

19. Kozuki N, Katz J, Lee ACC, et al. Short maternal stature increases risk of small for-gestational-age and preterm births in low and middle-income countries: Individual participant data meta-analysis and population attributable fraction. J Nutr. 2015;145(11):2542–2550. doi:10.3945/jn.115.216374

20. Patel A, Prakash AA, Das PK, Gupta S, Pusdekar YV, Hibberd PL. Maternal anemia and underweight as determinants of pregnancy outcomes: Cohort study in eastern rural Maharashtra, India. BMJ Open. 2018;8(8):1–15. doi:10.1136/bmjopen-2018-021623

21. Bhasin VK, Obeng C, Bentum-ennin I. Fertility, Income and Poverty of Households in Ghana. ResearchGate. Published online 2014. https://www.researchgate.net/publication/228416821

22. WHO. WHO Recommendations on Antenatal Care for a Positive Pregnancy Experience.; 2016. http://www.who.int

23. Al-Farsi YM, Brooks DR, Werler MM, Cabral HJ, Al-Shafei MA, Wallenburg HC. Effect of high parity on occurrence of anemia in pregnancy: A cohort study. BMC Pregnancy Childbirth. 2011;11(1):7. doi:10.1186/1471-2393-11-7

24. Cohen AK, Kazi C, Headen I, et al. Educational attainment and gestational weight gain among US mothers. 2017;26(4):460–467. doi:10.1016/j.whi.2016.05.009.

25. Educational Eggersdofer M, Kraemer K, Ruel M, et al. The road to good nutrition: A global perspective. In: Eggersdofer M, Kraemer K, Ruel M, et al., eds. Karger; 2013:27–30.

26. Zoet GA, Paauw ND, Groenhof K, et al. Association between parity and persistent weight gain at age 40-60 years: A longitudinal prospective cohort study. BMJ Open. 2019;9(5):1–8. doi:10.1136/bmjopen-2018-024279

27. Abrams B, Heggeseth B, Rehkopf D, Davis E. Parity and body mass index in US women: A prospective 25-year study. Obesity. 2013;21(8):1514–1518. doi:10.1002/oby.20503

28. United Nations Department of Economic Affairs Population Division. World Population Prospects: The 2006 Revision. Vol I. United Nations; 2007.

29. United Nations Department of Economic Affairs Population Devision 2015. Trends in Contraceptive Use Worldwide 2015. United Nations; 2015. doi:10.18356/f52491f9-en

30. Acharya O, Zotor FB, Chaudhary P, Deepak K, Amuna P, Ellahi B. Maternal Nutritional Status, Food Intake and Pregnancy Weight Gain in Nepal. J Health Manag. 2016;18(1):1–12. doi:10.1177/0972063415625537

31. Fanou-Fogny N, J. Saronga N Koreissi Y, A. M. Dossa R, Melse-Boonstra A, D. Brouwer I. Weight status and iron deficiency among urban Malian women of reproductive age. Br J Nutr. 2011;105(4):574–579. doi:10.1017/S0007114510003776

32. Assefa N, Berhane Y, Worku A. Wealth Status, Mid Upper Arm Circumference (MUAC) and Antenatal Care (ANC) Are Determinants for Low Birth Weight in Kersa, Ethiopia. PLoS One. 2012;7(6). doi:10.1371/journal.pone.0039957

33. WHO. Haemoglobin concentrations for the diagnosis of anaemia and assessment of severity. Geneva, Switz World Heal Organ. Published online 2011:1-6. doi:2011

34. WHO. Physical Status: The Use and Interpretation of Anthropometry. Vol 1995.; 1995.

35. Kushitor SB, Owusu L, Kushitor MK. The prevalence and correlates of the double burden of malnutrition among women in Ghana. PLoS One. 2020;15(12):1–12. doi:10.1371/journal.pone.0244362

36. Christian AK, Steiner-Asiedu M, Bentil HJ, et al. Co-Occurrence of Overweight/Obesity, Anemia and Micronutrient Deficiencies among Non-Pregnant Women of Reproductive Age in Ghana: Results from a Nationally Representative Survey. Nutrients. 2022;14(7). doi:10.3390/nu14071427

37. Cepeda-Lopez AC, Baye K. Obesity, iron deficiency and anaemia: A complex relationship. Public Health Nutr. 2022;23(10):1703–1704. doi:10.1017/S1368980019004981

38. Taghdir M, Alimohamadi Y, Sepandi M, Rezaianzadeh A, Abbaszadeh S, Mahmud FM. Association between parity and obesity: A cross sectional study on 6,447 Iranian females. J Prev Med Hyg. 2020;61(3):E476–E481. doi:10.15167/2421-4248/jpmh2020.61.3.1430

39. Rahman MA, Rahman MS, Aziz Rahman M, Szymlek-Gay EA, Uddin R, Islam SMS. Prevalence of and factors associated with anaemia in women of reproductive age in Bangladesh, Maldives and Nepal: Evidence from nationally-representative survey data. PLoS One. 2021;16(1):e0245335. doi:10.1371/journal.pone.0245335

40. Ausk KJ, Ioannou GN. Is obesity associated with anemia of chronic disease? A population-based study. Obesity. 2008;16(10):2356–2361. doi:10.1038/oby.2008.353

41. Qin Y, Melse-Boonstra A, Pan X, et al. Anemia in relation to body mass index and waist circumference among chinese women. Nutr J. 2013;12(1):10–12. doi:10.1186/1475-2891-12-10

42. Petry N, Wirth JP, Adu-Afarwuah S, et al. Risk factors for anaemia among Ghanaian women and children vary by population group and climate zone. Matern Child Nutr. 2021;17(2):1–10. doi:10.1111/mcn.13076

43. Liyew AM, Tesema GA, Alamneh TS, et al. Prevalence and determinants of anemia among pregnant women in East Africa; A multi-level analysis of recent Demographic and Health Surveys. PLoS One. 2021;16(4):e0250560. doi:10.1371/journal.pone.0250560

44. Teshale AB, Tesema GA, Worku MG, Yeshaw Y, Tessema ZT. Anemia and its associated factors among women of reproductive age in eastern Africa: A multilevel mixed-effects generalized linear model. PLoS One. 2020;15(9 September):1–16. doi:10.1371/journal.pone.0238957

45. Tirore LL, Mulugeta A, Belachew AB, et al. Factors associated with anaemia among women of reproductive age in Ethiopia: Multilevel ordinal logistic regression analysis. Matern Child Nutr. 2021;17(1):1–15. doi:10.1111/mcn.13063

46. Win HH, Ko MK. Geographical disparities and determinants of anaemia among women of reproductive age in Myanmar : analysis of the 2015 – 2016 Myanmar Demographic and Health Survey. who-seajph.org. 2018;7(September):107–113.

47. Pobee RA, Setorglo J, Klevor M, Murray-Kolb LE. The prevalence of anemia and iron deficiency among pregnant Ghanaian women, a longitudinal study. PLoS One. 2021;16(3 March):1–14. doi:10.1371/journal.pone.0248754

48. Fallatah AM, Babatin HM, Nassibi KM, Banweer MK, Fayoumi MN, Oraif AM. Maternal and Neonatal Outcomes among Obese Pregnant Women in King Abdulaziz University Hospital: A Retrospective Single-Center Medical Record Review. Med Arch (Sarajevo, Bosnia Herzegovina). 2019;73(6):425–432. doi:10.5455/medarh.2019.73.425-432

49. Rincón-Pabón D, González-Santamaría J, Urazán-Hernández Y. Prevalence and sociodemographic factors associated with iron deficiency anemia in pregnant women of Colombia (secondary analysis of the ENSIN 2010). Nutr Hosp. 2019;36(1):87–95. doi:10.20960/nh.1895

50. WHO. WHO Guideline on Use of Ferritin Concentrations to Assess Iron Status in Individuals and Populations. World Health Organization; 2020. https://www.who.int/publications-detail/9789240000124

51. Mwangi MN, Maskey S, Andang’o PEA, Noel KS, Johanna MR, Trijsburg L, et al. Diagnostic utility of zinc protoporphyrin to detect iron deficiency in Kenyan pregnant women. BMC Med. 2014;12(1). doi:10.1186/s12916-014-0229-8

